# Email vs. Instagram recruitment strategies for online survey research

**DOI:** 10.1101/2020.09.01.20186262

**Authors:** Rafael R. Moraes, Marcos B. Correa, Ândrea Daneris, Ana B. Queiroz, João P. Lopes, Giana S. Lima, Maximiliano S. Cenci, Otávio P. D’Avila, Claudio M. Pannuti, Tatiana Pereira-Cenci, Flávio F. Demarco

## Abstract

In this study, we describe and evaluate a method for reaching a target population (i.e., dentists practicing in Brazil) to engage in survey research using traditional e-mail invites compared with recruitment campaigns created on Instagram. A pre-tested questionnaire was used and participants were recruited for 10 days via a source list of email addresses and two discrete Instagram organic open campaigns. A total of 3,122 responses were collected: 509 participants were recruited by email (2.1% response rate) and 2,613 by the two Instagram campaigns (20.7% and 11.7% conversion rates), respectively. Response/min collection rates in the first 24 h ranged between 0.23 (email) and 1.09 (first campaign). In total, 98.8% of all responses were received in the first 48 h for the different recruitment strategies. There were significant differences for all demographic variables (p<0.001) between email and Instagram respondents, except for sex (p=0.373). Instagram respondents were slightly older, had more professional experience (years in practice), and a higher graduate education level than email respondents. Moreover, most email and Instagram respondents worked in the public sector and private practice, respectively. Although both strategies could collect responses from all Brazilian regions, email responses were slightly better distributed across the five territorial areas compared to Instagram. In conclusion, this study provides evidence that survey recruitment of a large sample using Instagram is feasible. However, using Instagram to engage participants is challenging and has limitations that warrant further investigation. Combination of email and Instagram recruitment led to a more diverse population and improved response rates.

## 1. INTRODUCTION

A major challenge in survey research is developing a method for recruiting potential participants that reaches a broad respondent range and allows a sample with diverse characteristics that is representative of the target population. Traditional surveying strategies, including in-person and telephone recruitments, are time-intensive but can yield good response rates [1-4]. Recruitment via the Internet became frequent when emails were the most used online media and still remains a popular method. The major drawbacks of email recruitment include the difficulty in gathering updated email addresses [5] and fluctuating response rates [2,3,6].

Social networking services are currently used in daily communications and can be considered in important learning and collaboration developments in healthcare [7,8]. Facebook, YouTube, and Instagram are currently the top three most popular social networks [9]. Instagram is a photo-and video-sharing service that allows users to make visual and textual meanings to interact with ambient viewers [10]. It is currently the fastest growing social media platform and reached 1 billion monthly active users worldwide in 2020 [9]. Instagram is among the most frequently used tools by the World Health Organization and other public health agencies for disseminating visually rich health-related messages [8,11]. Current use of Instagram as a research tool can be categorized into educational/informational purposes and motivational/supportive applications [8].

Facebook has been used in research to reach target populations [3,12-17]. Contrastingly, there have been few reports regarding the use of Instagram for the same purpose. A recent study [13] showed that Facebook and Instagram audience-tailored strategies allowed the recruitment of a large sample of a population characterized as hard-to-reach (i.e., LGBT+ young adults). Another study [15] reported that paid advertising on Facebook, Instagram, and Snapchat promoted the recruitment of youths to a prevention campaign. There have been no reports regarding the use of Instagram as a method for engaging health professionals in survey research. A significant challenge is that website links are not allowed in Instagram posts and comments, which increases the number of steps required for potential participants to reach the online collection tool. However, the homophilic character of online social networks may favorably contribute to their use as recruitment methods since individuals tend to associate, follow, and bond with similar individuals [18]. Novel online surveying/recruitment strategies are especially important for situations such as the coronavirus disease (COVID-19) pandemic where sanitary measures prevent traditional research approaches. Specifically, the pandemic may facilitate online recruitment since people have been spending more time at home and on social media [19]. This research type may be largely used in the next years when there is a risk of a COVID-19 resurgence [20]. This is further reinforced by the potential better cost-effectiveness of social media recruitment compared with traditional enrollment methods [17].

However, the use of social media as a research tool presents new challenges to those already present in traditional surveying strategies, including the expected sociodemographic diversity from different recruitment approaches and the generalizability of research findings [3,21]. This study aimed to describe a method for reaching a target population (i.e., dentists) to engage in survey research via traditional e-mail invites combined with a recruitment campaign created on Instagram. Specifically, this study aimed to determine the feasibility of using Instagram to recruit survey participants and analyze respondents reached by Instagram and email, as well as relevant methodological aspects related to this study design.

## 2. METHODS

To illustrate this methodological paper, an online survey targeting dentists in Brazil is described, with a focus on the recruiting strategy rather than the participants. We have previously attempted to reach dentists working in Brazil via emails sent by regional dental councils with low return rates [6]. Therefore, in a recent study addressing the COVID-19 impact on dentistry [22], we prospected to additionally use Instagram to reach our target population since it is among the most popular social networks worldwide. Further, it is widely used by dentists in Brazil, with #dentistry and #odontologia (Portuguese for dentistry) having been used in 5.4M and 7.3M posts by September, 2020, respectively. To our knowledge, this was the first study to use Instagram to reach healthcare professionals to engage in survey research [22].

### 2.1. Study design

Dentists were the target population of the open survey. Brazil has the largest population of dental professionals in the world [23], with more than 348,000 dentists working in the public network and/or in private offices. The study protocol was approved by our institutional review board (#4.015.536). A 30-item questionnaire (three screens) was developed, pre-tested, and used in a cross-sectional survey. The questionnaire items mainly assessed the impact of COVID-19 on the dental practice routine [22]; however, the scope of the present report does not include COVID-19 and pandemic-related questions. In accordance with open science practices, the research protocol, questionnaire in its original language, databank of responses, and other study-related information have been published in an open platform (doi:10.17605/OSF.IO/DNBGS). An English translation of the questionnaire is provided in the Appendix. The Checklist for Reporting Results of Internet E-Surveys (CHERRIES) [24] and The SUrvey Reporting GuidelinE (SURGE) [25] were consulted for this report. A checklist of CHERRIES items is provided in the Appendix.

### 2.2. Questionnaire development and pre-testing

The study protocol was detailed in our previous report [22]. A self-administered electronic questionnaire was developed based on the inputs of at least 8 researchers in three discrete review rounds. The questionnaire was hosted online on Google Forms (Google Inc., Mountain View, CA, USA) and pre-tested using a sample of 22 dentists from different locations in Brazil. The dentists were asked to evaluate clarity, writing style, question sequence, and internal consistency. This allowed the assessment of the reliability and face validity of the tool and items. There were differences in sex, age, working sector, country region, experience, and education levels across the pre-testers to represent the population of dentists in Brazil. The pre-testers were first asked to respond to the questionnaire and record the time taken to complete it: the average time ± standard deviation (SD) was 7 ± 2 min. In the second series, the pre-testers ranked the clarity of each question using scores between 1 (not clear) and 5 (very clear) on a Likert scale. There was a text box after every question for pre-testers to explain their scores and place comments, critics, and suggestions, including other response options. Questions rated with scores ≤ 3 (n = 9) were discussed by at least three researchers and edited based on the pre-tester comments. The average ± SD scores were 4.79 ± 0.10 and 4.91 ± 0.11 for the 9 questions requiring revision and all 30 questions, respectively. The pretest was important to include other response options, which aided in reducing response bias. The questionnaire was reviewed and revised iteratively by the executive group for approval. Pre-testers were excluded from the definitive study.

### 2.3. Questionnaire content

The original language was Brazilian Portuguese; however, the questionnaire was translated into English and is presented in the Appendix. The first page of the questionnaire contained the study title and objective, as well as an invitation exclusively for dentists to participate and complete the questionnaire only once. They were informed that their participation was voluntary and not paid; moreover, they were informed regarding the potential risks and benefits, as well as were assured that all responses would be treated confidentially and anonymously. In addition, the respondents were asked not to complete the questionnaire if they were not dentists or they had previously responded, which reduced the risk for duplicate answers. No other means to prevent multiple entries were used. The participants were asked to print or save the first page of the questionnaire as a PDF file to retain a copy of the informed consent form. Contact information of the researchers and institutions responsible for the survey was provided. The participant had to click ‘Yes’ after the question “Do you agree to participate in the study voluntarily?” to access the questionnaire. By clicking ‘No’, the survey was terminated. The questionnaire contained 30 mandatory close-ended questions; among them, 8 were related to the demographic and professional profile of the respondents and have been addressed in the present report. The options ‘I’d rather not say’, ‘I don’t know how to answer’, and ‘Does not apply’ were available for all questions to avoid response errors and to reduce possible discomfort in answering any question. Open-ended questions were not used to avoid the need for extensive data review, except for the option “other” in 9 questions. The main outcomes were related to the professionals’ behavior regarding their practice routines before and during the pandemic; however, aspects regarding the pandemic are beyond the scope of this article.

### 2.4. Participant recruitment via e-mail

The participants were recruited for 10 days. The first strategy for reaching out to the target population was the use of traditional email invites. A source list of 24,126 emails of dentists who mainly work in the public healthcare network was used. An online survey software was used to send the email invites (SurveyMonkey, San Mateo, CA, USA). The email body contained a short text regarding the study objective, average response time, and the university conducting the study. The online software did not allow placement of the Google Form questionnaire link on the email body. Therefore, the participants had to undergo a two-stage process to access it: clicking on the ‘begging the questionnaire’ button on the email body and then clicking on the ‘access the questionnaire’ link on a second screen. The first email was sent on May 15, 2020 and a reminder email was sent after five days.

### 2.5. Participant recruitment via Instagram

A second recruitment strategy was the creation of a campaign directed to dentists on Instagram. The campaign was created in Portuguese and restricted to organic reach among Instagram users, that is, there was no paid advertising. The campaign started on May 20, 2020, which was the date the reminder emails were sent. Since the participants were not asked to identify themselves, starting the Instagram campaign later was important to secure an exclusive period for email responses. The campaign indicated that dentists were invited to participate in an online survey regarding the impact of the pandemic on their routine practice. An Instagram professional account was created (@odcovid) with a short username to facilitate search. Ads were created on PowerPoint (Figure 1), exported into PNG image files, and posted on @odcovid feed and stories. In the post captions, we used hashtags related to dentistry and COVID-19 to increase the reach of the target population. The advertisements called for the attention and participation of dentists with brief descriptions of the study objective, average response time, and the university conducting the study; moreover, the questionnaire website link was available on the @odcovid bio page. This was necessary because Instagram does not allow website links in posts or captions. Thus, participants recruited via Instagram were required to complete a two-stage process to access the questionnaire: search for @odcovid profile in the app and click on the weblink available on the bio page. An Instagram bio is the initial screen when one accesses a profile. It is found underneath the username and contains a small summary of the profile, including contact information and a website link.

**Figure 1.**
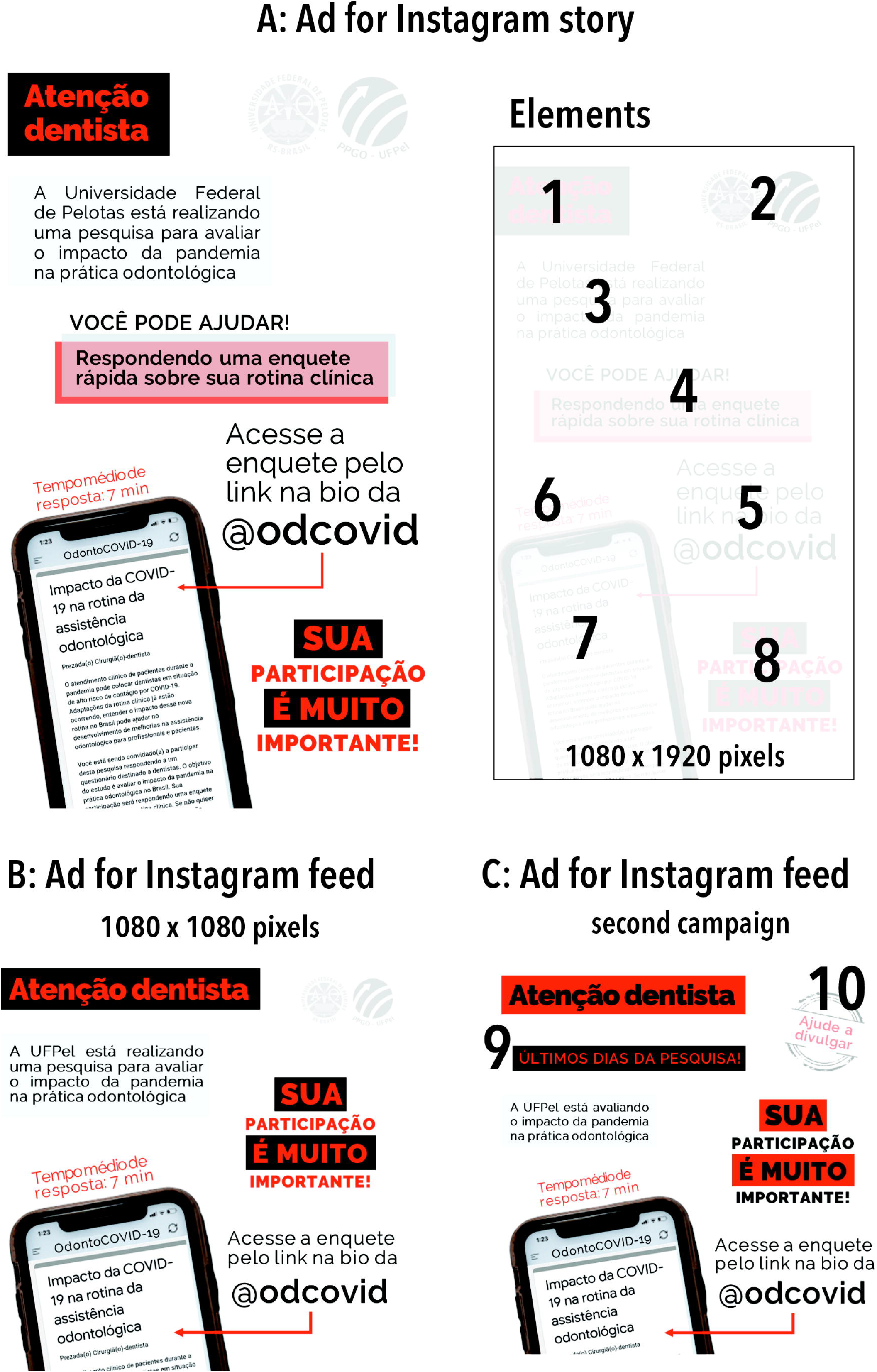
Original ads used to recruit dentists on Instagram with aspect ratios shown in pixels. A: Ad for Instagram story. The marked elements are: 1) mention that the call was for dentists; logos of the institution conducting the study; 3) informative short text indicating the university conducting the research and the study objective; 4) indication that participation would involve response to a questionnaire; 5) information that a website link to the questionnaire was available at @odcovid; 6) average response time (7 min) collected at the pre-test; 7) a cell phone containing the first page of the questionnaire was shown; 8) a statement reinforcing that participation was very important. B: Ad for Instagram feed, which had fewer elements than A given its different size and aspect ratio compared with the ad for Instagram story. All elements presented in A were used in B with slight edits, except for element 4. C: Ad with similar content, but a slightly different look, as B, which also indicated that research was in the final days of response collection (9) and asked for help in sharing the survey (10) to aid in creating a snowball sampling in the social media. The images were prepared in PowerPoint to have an aesthetically pleasing appearance but to keep a sober tone. Font size varied between 24 (element 6) and 88 (@odcovid in element 5).

Moreover, the participating researchers posted the advertisements on their personal Instagram profiles (feed and/or stories) on the day the campaign was started. To expand the promotion, Brazilian dentists with professional Instagram profiles were asked to disseminate the advertisements with survey invitations. We sent messages via Instagram Direct containing the images used in feed and stories posts to dentists who agreed to repost the advertisements. At least 50 Instagram accounts, which were categorized as either micro (< 10,000 followers) or meso (10,000–1 million) follower scales [26], were asked to repost the first campaign ads. A second Instagram campaign with similar content, but with a slightly different look (Figure 1C), was created after two days later using a similar methodology except for the inclusion of a sentence that asked the audience to help in disseminating the campaign themselves, which could aid in creating snowball sampling in the social network. Different Instagram users (at least 25 dentists from different Brazilian locations) were asked to repost the advertisements during the second campaign.

### 2.6. Sample selection and size, survey administration, and collection of responses

All dentists practicing in Brazil were eligible for study participation. The pathway to reach the questionnaire differed between the email and Instagram approaches; however, they both involved a two-stage process, as aforementioned. Survey administration was the same since both pathways led the respondents to a unique website link. When the participants clicked on the weblinks provided in the email or the Instagram bio page of @odcovid, they accessed the questionnaire in Google Forms. We tested whether the questionnaire could be read well in different computers, tablets, and cell phones prior to the start of the campaign. No randomization of items or questionnaires or adaptive questioning were used. Based on a population of ~348,000 dentists in Brazil, we estimated that 2,385 responses were required to ensure a 95% confidence interval with a 2% margin of error. Responses were collected between May 15 and 24, 2020. The timeframe for response collection was not long since the turbulent pandemic scenario could have led to significant changes in the dentists’ behavior in the short term.

### 2.7. Data analysis

Partial questionnaire completion was impossible and review was allowed until submitting responses. ‘I’d rather not say’, ‘I don’t know how to answer’, and ‘Does not apply’ responses were treated as missing data; therefore, the sample size varied among different responses. No questionnaires submitted with an atypical timestamp were observed. The number of responses and response/min collection rates in the first 48 h of recruitment were calculated for each approach. The following numerical outcomes were collected on an Instagram insights dashboard for both campaigns created on @odcovid: likes, direct message shares, comments, unique accounts reached, impressions (i.e., number of times that the post was viewed), website clicks, and story shares. The conversion rates were calculated, that is, the proportion of impressions that led to website clicks. Descriptive statistics were used to identify the frequencies and distributions of variables between the email vs. Instagram respondents. Proportions were compared using chi-square tests. Some data presented here have been previously reported [22].

## 3. RESULTS

### 3.1. Data on responses received via different recruitment strategies

A total of 3,122 valid responses were collected over 10 days, with the dates and times of the responses being recorded on Google Forms. A total of 37 participants clicked on the ‘no’ response, which indicated that they declined to participate after reading the first questionnaire page (1.2% refusal rate, 98.8% completion rate). Figure 2 shows the gathering of valid responses over time and important research timepoints. The response collection dynamics may be divided into three main recruitment phases:

- <u>Phase 1 (day 1 through 5):</u> In periods A–B shown in Fig.2, responses were exclusively received via e-mail invites sent on day 1; thus, the population in this phase only included email respondents. A total of 509 responses were received over five days, which comprised an email response rate of 2.1%. The number of actual rejections/losses could not be calculated because we could not estimate the number of dentists who received the email invites or decided not to respond to the questionnaire, as well as the number of emails that went to spam folders. Unique site or survey visitors were not registered.
- <u>Phase 2 (day 6 through 8):</u> The first Instagram campaign started on day 6 (B in Fig. 2); subsequently, on the same day, a reminder email was sent to all individuals in the source email list (C in Fig.2). A total of 1991 responses were received in this phase. The response rates in Instagram could not be estimated; however, conversion rates were calculated, which may provide an estimation of return rates, as later detailed in this section. Responses received in Phase 2 mainly originated from Instagram, which was indicated by the remarkably higher response number compared with Phase 1. However, we acknowledge that some email responses could have been present. Thus, Phase 2 could be described as a mixed recruitment phase that was largely determined by Instagram respondents.
- <u>Phase 3 (day 8 through 10):</u> Two days after starting the second Instagram campaign (D in Fig.2), collection of responses was terminated. During this period, 622 responses were received (return rates are described below). This phase almost exclusively contained Instagram responses; however, some late email responses could also have been received. Phase 3 can be described as involving recruitment exclusive via Instagram.

**Figure 2.**
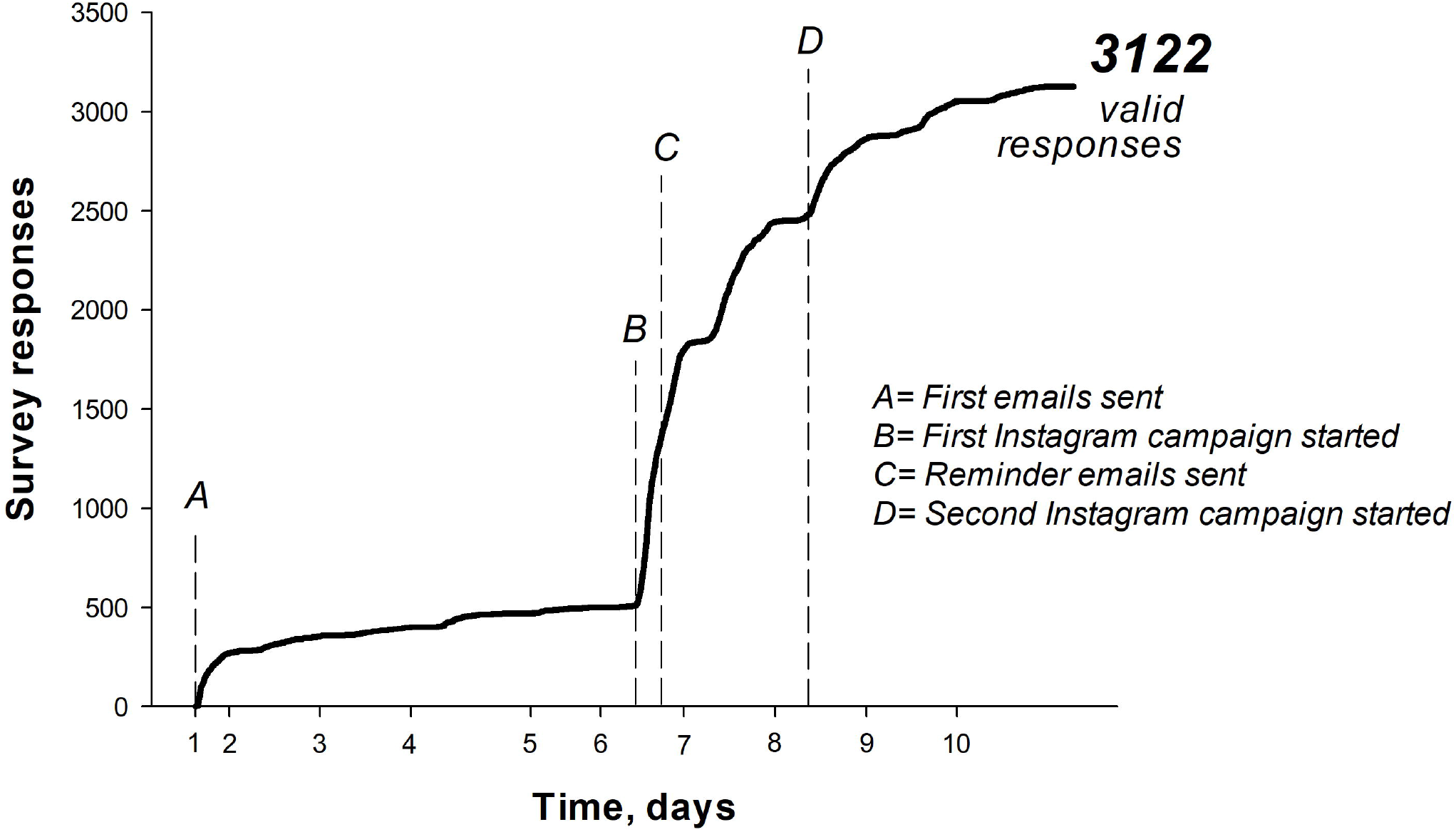
Collected survey responses against time (days). Letters A–D indicate different timepoints of email invites and Instagram campaigns. In 10 days, 3,122 valid responses were received.

Table 1 presents the number of responses and response/min collection rates during the first 48 h of recruitment using different strategies. In the first 24 h of each phase, 325, 1572, and 557 responses were collected in Phase 1, 2, and 3, respectively. The response/min collection rates ranged between 0.23 (email) and 1.09 (first Instagram campaign). Between 24 h and 48 h of recruitment, responses for email and the first Instagram campaign reduced by 82.5% and 67.8%, respectively, compared with the first 24 h. The 24–48 h response/min rates were lower for email invites than for the first Instagram campaign; however, they were closer to the values for the second Instagram campaign. Based on the number of responses received in the first 48 h for the three different recruitment phases, the total number of responses was 3086, which translated to 98.8% of all responses.

**Table 1.** Number of responses and response/min rates during the first 48 h for the different recruitment approaches (N = 3,122)

|  | Responses, n (% of total) |  |  | Response/min collection rate |  |
| --- | --- | --- | --- | --- | --- |
|  | First 24 h | 24–48 h | Reduction* | First 24 h | 24–48 h |
| Phase 1: First email invites | 325 (10.4) | 57 (1.8) | 82.5% | 0.23 | 0.04 |
| Phase 2: First Instagram campaign + email reminder | 1,572 (50.3) | 506 (16.2) | 67.8% | 1.09 | 0.35 |
| Phase 3: Second Instagram campaign | 557 (17.8) | 69 (2.2) | 87.6% | 0.39 | 0.05 |

\*Difference between 'first 24 h' and '24–48 h' periods.

Table 2 presents Instagram data insights for the two recruitment campaigns. The number of likes, shares, comments, and unique accounts reached was lower during the second campaign. The conversion rate was 20.7% and 11.7% in the first and second campaigns, respectively. A total of 442 questionnaire website clicks originated from these two Instagram feed posts, which suggested that at least 2171 Instagram respondents accessed the questionnaire by searching @odcovid in the app.

**Table 2.**
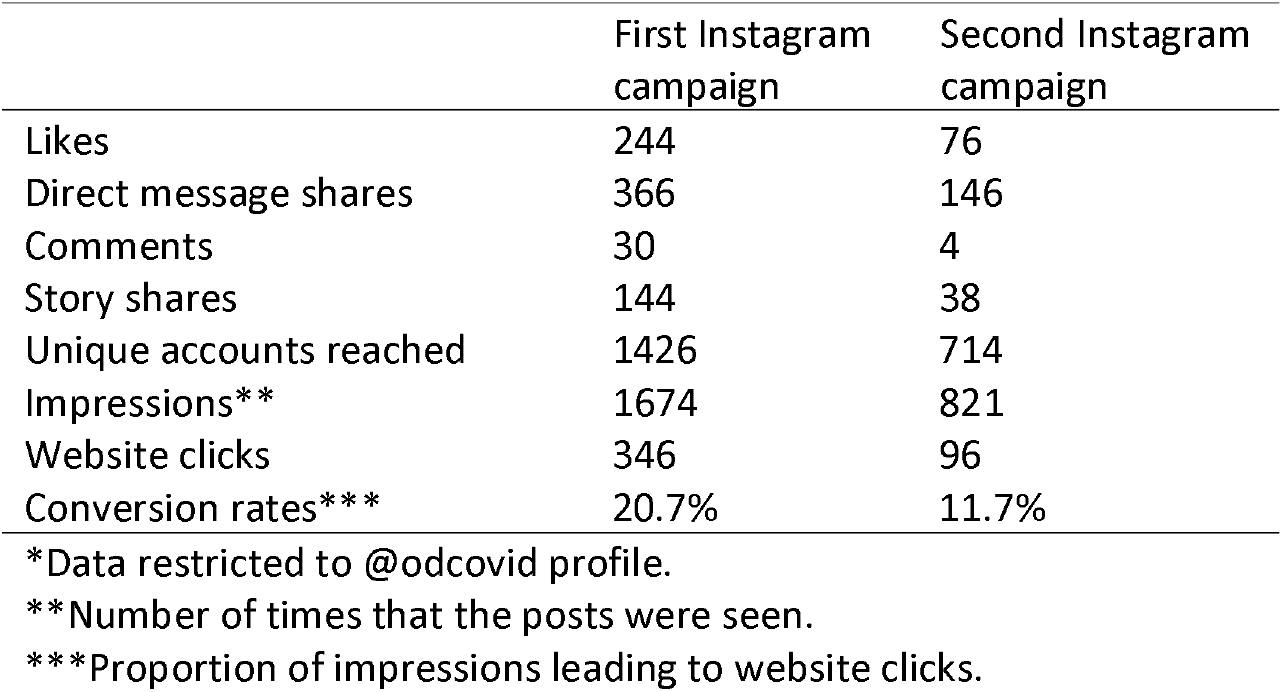
Data insights for the posts on Instagram feed* in the discrete campaigns

|  | First Instagram campaign | Second Instagram campaign |
| --- | --- | --- |
| Likes | 244 | 76 |
| Direct message shares | 366 | 146 |
| Comments | 30 | 4 |
| Story shares | 144 | 38 |
| Unique accounts reached | 1426 | 714 |
| Impressions** | 1674 | 821 |
| Website clicks | 346 | 96 |
| Conversion rates*** | 20.7% | 11.7% |

\*Data restricted to @odcovid profile.
\*\*Number of times that the posts were seen.
\*\*\*Proportion of impressions leading to website clicks.

Comparisons of populations among Phases 1, 2, and 3 revealed significant differences for all demographic and work variables (p < 0.0001) except for sex (p = 0.157). Comparison of Phases 2 and 3 with Phase 1 revealed differences only for changes in frequencies. Therefore, since most responses in Phases 2 and 3 were collected via Instagram recruitment, we decided to report the results with consideration of only two distinct populations: email respondents (Phase 1) and Instagram respondents (Phases 2 + 3).

### 3.2. Overall characteristics of the respondents

Responses were received from all 26 Brazilian states and the federal district. Table 3 presents the demographic and work practice characteristics of the dentists who responded to the questionnaire. As aforementioned, the sample size varied among different variables due to missing data. Most responses were received after the first Instagram campaign started (83.7%). The respondents were mostly females (74.7%) and had been in practice for up to 20 years (73.9%). The mean age ± SD of the respondents was 38 ± 11 years. The X and Y generations accounted for 93% of participants. Residency or advanced special training was the most often reported graduate education level (49%). Meanwhile, 53% and 36% of dentists worked mainly in private clinics and the public sector, respectively. Dentists from all Brazilian regions responded to the questionnaire with a predominance of responses from the South and Southeast regions (67.4%).

**Table 3.** Demographic and work practice characteristics of the respondents (N = 3,122)

| Variable/category | n* | % | 95% CI |
| --- | --- | --- | --- |
| **Recruitment method** | 3122 |  |  |
| Email | 509 | 16.3 | 15.0; 17.6 |
| Instagram | 2613 | 83.7 | 82.4; 85.0 |
| **Sex** | 3,116 |  |  |
| Male | 790 | 25.4 | 23.9; 26.2 |
| Female | 2,326 | 74.7 | 73.1; 76.2 |
| **Generation** | 3,121 |  |  |
| Baby boomers | 227 | 7.3 | 6.4; 8.3 |
| X | 1,311 | 42.0 | 40.3; 43.7 |
| Y | 1,593 | 50.7 | 48.9; 52.5 |
| **Years in practice** | 3,121 |  |  |
| ≤ 10 | 1,496 | 47.9 | 46.2; 49.7 |
| 11–20 | 812 | 26.0 | 24.5; 27.6 |
| 21–30 | 501 | 16.1 | 14.8; 17.4 |
| > 30 | 312 | 10.0 | 9.0; 11.1 |
| **Graduate education** | 3,121 |  |  |
| None | 758 | 24.3 | 22.8; 25.8 |
| Residency or advanced special training | 1,530 | 49.0 | 47.3; 50.8 |
| MSc or PhD | 833 | 26.7 | 25.2; 28.3 |
| **Main practice sector** | 3,051 |  |  |
| Public | 1,091 | 35.8 | 34.1; 37.5 |
| Private | 1,601 | 52.5 | 50.7; 54.2 |
| Other | 359 | 11.8 | 10.7; 13.0 |
| **Brazilian region** | 3,122 |  |  |
| South | 1,183 | 37.8 | 36.2; 39.6 |
| Southeast | 923 | 29.6 | 28.0; 31.2 |
| Central-west | 221 | 7.1 | 6.2; 8.0 |
| Northeast | 682 | 21.9 | 20.4; 23.3 |
| North | 113 | 3.6 | 3.0; 4.3 |

\*Varies from total N because of missing data for different questions.

### 3.3. Email vs. Instagram populations

Figure 3 presents a comparison of the frequencies of responses received by email vs. Instagram recruitment strategies. There were significant differences for all variables (p < 0.001) except for sex, which was similar between email and Instagram (p = 0.373). Email respondents were younger and generally had less professional experience (years in practice as dentists) than Instagram respondents. Instagram respondents reported higher levels of graduate education (MSc or PhD degrees) than email respondents, who more often reported not having completed any graduate education. Professionals working in the public sector were mainly reached by email; however, some of the Instagram respondents were public practicing dentists. The distribution of respondents by territorial areas showed that both email and Instagram could recruit respondents from all regions; however, the combination of these two recruitment strategies improved the distribution of responses across Brazilian regions.

**Figure 3.**
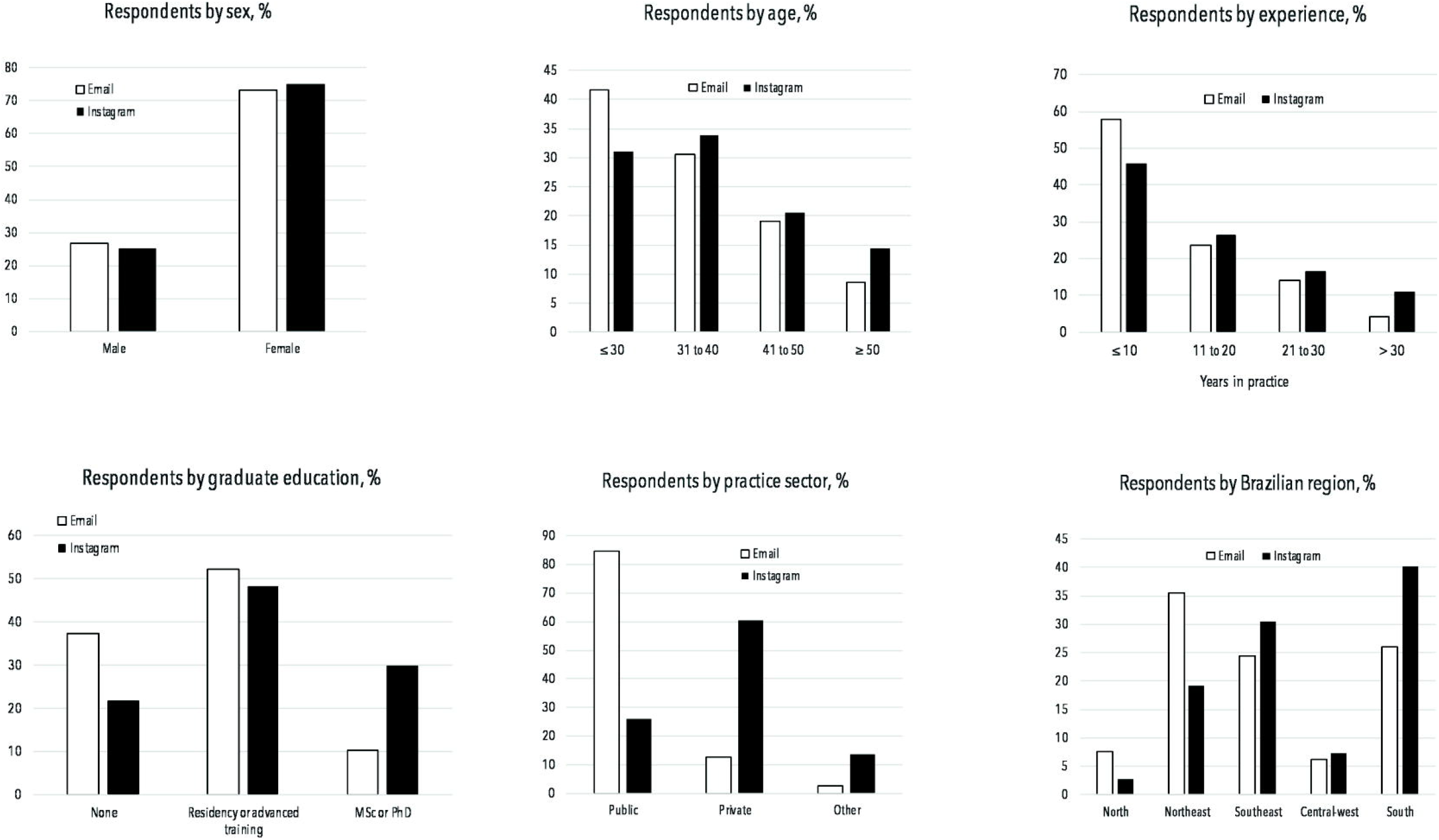
Frequencies of responses received by different recruitment strategies: email vs. Instagram. There were no significant differences in sex between populations recruited by different strategies (p = 0.373), whereas all other variables were significantly different between email and Instagram respondents (p < 0.001).

Table 4 presents a comparison of demographic data of dentists working in Brazil compared with the distribution of responses according to sex, age, and Brazilian region. Compared with the general dentist population, there was a larger predominance of females among both email and Instagram recruitments. Responses were well distributed by age as compared with the general population, especially for Instagram respondents. In contrast, email responses were slightly better distributed across the five territorial areas than Instagram responses.

**Table 4.** Distribution of dentists working in Brazil by sex, age, and region (%) compared with the survey participants recruited by different strategies

| Variable/category | Dentists working in Brazil* | Survey respondents |  |
| --- | --- | --- | --- |
|  |  | Email | Instagram |
| **Sex** |  |  |  |
| Male | 43.9 | 26.9 | 25.1 |
| Female | 56.1 | 73.1 | 75.0 |
| **Age** |  |  |  |
| ≤ 30 | 25.2 | 41.6 | 31.0 |
| 31–40 | 32.2 | 30.6 | 33.9 |
| 41–50 | 23.6 | 19.1 | 20.1 |
| 51–60 | 14.1 | 8.1 | 10.3 |
| > 60 | 4.9 | 0.6 | 4.1 |
| **Brazilian region** |  |  |  |
| South | 16.1 | 26.0 | 40.2 |
| Southeast | 52.8 | 24.4 | 30.6 |
| Central-west | 8.8 | 6.3 | 7.2 |
| Northeast | 16.6 | 35.6 | 19.2 |
| North | 5.7 | 7.7 | 2.8 |

\*Data obtained from official reports [23,27].

## 4. DISCUSSION

This is one of the first studies to use Instagram to reach a target population for engaging in survey research. To our knowledge, this is the first study to compare the characteristics of populations recruited via email vs. Instagram. Generally, Instagram respondents were slightly older, had more professional experience (years in dentistry practice), and a higher graduate education level than email respondents. Moreover, most email and Instagram respondents worked in the public sector and private practice, respectively. Although both strategies could collect responses from all Brazilian regions, email responses were slightly better distributed across the five territorial areas compared to Instagram. Instagram had a higher frequency of respondents from the South, which was the region where the social network campaign started. Therefore, it seems that combining email and Instagram recruitment strategies is important for increasing sample diversity. These findings are important for survey research using social media recruitment, which is a method likely to be more common in the future given the COVID-19 pandemic scenario. A recent study reported that determining the profile of the target sample is important for choosing the online recruitment method [4].

Instagram recruitment collected more than five times the number of responses collected by email. However, one must consider that the number of emails in the list was likely much lower than the number of dentists who potentially saw the Instagram recruitment campaigns. The second Instagram campaign was less effective than the first one in engaging participants but still yielded a similar response/min collection rate to emails. To explain the relative success of this completely ‘organic’ campaign in engaging participants to respond to a questionnaire, one should consider different aspects. First, the target population was large: Brazil has the largest population of dentists in the world, who use Instagram very often. Second, this was the first campaign on Instagram for dentists, and probably any healthcare area, in Brazil; thus, novelty could have attracted attention. Soon after the campaign was created, a series of similar surveys with dentists, dental students, and other healthcare professionals in Brazil were posted on Instagram. Until August 2020, we were able to count 21 similar recruitment campaigns. Moreover, in May 2020, the COVID-19 epidemic curve showed early signs of escalation and the Brazilian Ministry of Health indicated that dental appointments should be restricted to urgencies. Dentists (and general people) were staying more at home and using social media more often, and thus were more likely to be engaged. The pandemic burden to dental professions might have also favored recruitment since participants felt that they were contributing to their dental sector during difficult times.

The relative success of our recruitment strategy on Instagram could also be attributed to how people interact in social networks. The diffusion of information is largely determined by homophily, that is, a tendency for similar people to be connected [28,29]. This means that people interact with other individuals with similar ideologies, backgrounds, behaviors, and/or interests. A recent study showed that introducing a positive algorithmic bias could not stop online segregation in social networking [30]. Targeting a specific population has been reported as important for increasing e-mail survey response rates [31]. However, recruiters might have an additional role in online social media surveys. Since dental professionals posted the ads with dentists as targets, the connections were probably well established given the social and work similarities between recruiters and participants. Social media networking is also driven by the building of reputation, social capital, reciprocity, altruism, and trust [32], which likely breeds more social connections than email invites. Furthermore, these aspects may explain why more Instagram responses were received by participants from the region where the survey began. In addition, the present results are suggestive of the importance of reposts by users from different regions in promotions across large territorial areas, at least in cases where organic reach is used for recruitment.

Another relevant finding was that 98.8% of all responses were gathered in the first 48 h after initiation of each recruitment strategy, that is, after the first emails were sent or the discrete Instagram campaigns were created. This suggests that leaving an online questionnaire open for longer than two days may not greatly benefit participant recruitment. This is more likely the case for Instagram, where posts have a limited diffusion time; however, email responses were also concentrated within the first few days. Therefore, sending email reminders or reposting ads on social media is encouraged after 48 h. A shortcoming of Instagram campaigns is that response rates cannot be calculated. An alternative is to calculate the conversion rates (Table 2). Assuming that all website clicks at the @odcovid bio page link led to a valid response, only 16.9% of respondents would have been recruited by the ads posted by @odcovid. Although we could not calculate numbers, the campaigns could have reached a very large population, which allowed recruitment of the remaining 83.1% of the respondents. Since website clicks do not necessarily lead to survey responses, it is possible that the proportion of respondents recruited by reposts was even higher. This finding confirms the importance of other Instagram users reposting advertisements. Although the questionnaire link was only available on Instagram, the participants may have sent the link to colleagues in other ways, for example, direct messages on Instagram or other social media. Therefore, for surveys recruiting participants through open social media campaigns, researchers are encouraged to include a question in the questionnaire regarding how the respondents became aware of the recruitment.

Notably, email respondents were younger and had lower graduate education levels compared with Instagram respondents. There is a need for further studies to confirm these findings and identify the factors underlying these results. A study in the same region reported that Facebook was the most effective method for recruiting young adults [4]. Our results could have been influenced by the characteristics of the population in the convenience email sample used as a source list. For example, a higher prevalence of less experienced dentists could be a factor. Moreover, the respondents’ age and familiarity with mobile tools could affect their readiness to check emails and affect response rates since younger people may own and use more cell phones than older people who are relatively passive cell phone users [33]. However, in the present study, both recruitment methods were performed online. In addition, a previous study reported no major differences between older and younger users when physically interacting with digital assistants to complete tasks, including pressing buttons, viewing icons, recording messages, and scanning bar codes [34].

In surveys, it might be important to determine whether the sampling method can recruit a population that is representative of the target population. In the present study, representativeness may not be a factor since the study objective was to discuss methodological aspects and compare respondent samples. However, data shown in Table 4 provide insights regarding whether the dentists recruited by email and Instagram show similarities with the general population of dentists in Brazil. There were relative similarities between the characteristics of the general practitioners and those of the samples, apart from the aforementioned younger age and less experience among the email respondents. Notably, there was a higher response rate among Instagram respondents from Southern Brazil, which differed with the general distribution of dentists. The use of social media campaigns for survey recruitment seems powerful; however, the potential sampling bias should not be underestimated [21]; specifically, with respect to collecting more responses from regions near where the campaign began. Therefore, there is a need to determine the means for improving recruitment methods targeting respondents from far regions.

The combination of email and Instagram recruitment seemed to lead to a more diverse population and improve response rates, which is consistent with previous studies involving a combination of different recruitment methods [1,3,4,35]. Future studies could assess the reasons for survey participation based on the different types of recruitment approaches. In the present study, email and Instagram recruitments followed a very similar approach; however, the emails only had text and lacked pictures, which may lead to differences in communication channels and influence response rates [36]. In addition, social media posts might be seen multiple times by participants; contrastingly, although emails are an exclusive invitation, they may not be seen as often. However, these differences between email and Instagram are not expected to introduce measurement differences similar to those observed between face-to-face and internet surveys where the presence or lack of personal contact, as well as the associated social factors, may lead to distinct answer patterns [37]. To improve recruitment, other ways to motivate participants may be used, including giveaways and monetary incentives [35,38]. The effect of paid advertising on recruitment could also be investigated since social media organic reach is likely to continue shrinking given that social networks are placing strategies for monetizing platform investments.

## 5. CONCLUSIONS

The present study provides evidence that survey recruitment of a large sample using Instagram is feasible and that effective recruitment can be done in the short term. However, using Instagram to engage participants in survey research is challenging and has limitations that warrant further investigation. Although the cost-effectiveness of social media campaigns is attractive and using Instagram could recruit at least five times the number of participants recruited by email, our findings suggest that combining email and Instagram recruitment can lead to a more diverse population and improve response rates.

## Data Availability

The Checklist for Reporting Results of Internet E-Surveys (CHERRIES) and The SUrvey Reporting GuidelinE (SURGE) were consulted for this report. A checklist of CHERRIES items is provided in the Appendix.

https://osf.io/dnbgs/

## ACKNOWLEDGEMENTS

We also thank all persons who helped to disseminate the campaign on Instagram and dentists who volunteered to participate.

## AUTHOR CONTRIBUTIONS

RRM: contributed to conceptualization, investigation, methodology, validation, data curation, supervision, funding acquisition, project administration, writing, editing, and critical review of the manuscript. MBC: contributed to conceptualization, investigation, methodology, validation, data curation, software, formal analysis, writing, and critical review. AD, ABQ, JPL: contributed to methodology, validation, data curation and translation, and proofreading of the manuscript. GSL: contributed to conceptualization, methodology, data curation, writing, and critical review. MSC: contributed to conceptualization, methodology, validation, writing, and critical review. CMP: contributed to data curation, writing, critical review, and proofreading of the manuscript. OPD: contributed to conceptualization, methodology, and critical review. TPC: contributed to conceptualization, methodology, and critical review. FFD: contributed to conceptualization, supervision, funding acquisition, writing, and critical review. Order of authors considered relative contribution and gender equality. All authors gave their final approval and agree to be accountable for all aspects of the work.

